# Predicting Clinical Dementia Rating Using Blood RNA Levels

**DOI:** 10.1101/19009092

**Authors:** Justin B. Miller, John S.K. Kauwe, for the Alzheimer’s Disease Neuroimaging Initiative

**Affiliations:** Department of Biology, Brigham Young University, Provo, UT 84602, USA

**Keywords:** Diagnosis, machine learning, CLIC1, Clinical Dementia Rating

## Abstract

**INTRODUCTION:** The Clinical Dementia Rating (CDR) is commonly used to assess cognitive decline in Alzheimer’s disease patients.

**METHODS:** We divided 741 participants with blood microarray data in the Alzheimer’s Disease Neuroimaging Initiative (ADNI) into three groups based on their most recent CDR assessment: cognitive normal (CDR=0), mild dementia (CDR=0.5), and probable AD (CDR≥1.0). We then used machine learning to predict cognitive status using only blood RNA levels.

**RESULTS:** One chloride intracellular channel 1 (CLIC1) probe was significant. By combining nonsignificant probes with p-values less than 0.1, we averaged 87.87 (s = 1.02)% predictive accuracy in classifying the three groups, compared to a 55.46% baseline for this study.

**DISCUSSION:** We identified one significant probe in CLIC1. However, CLIC1 levels alone were not sufficient to determine dementia status. We propose that combining individually suggestive, but nonsignificant, blood RNA levels can significantly improve diagnostic accuracy.

## Background

Late-onset Alzheimer’s disease (AD) permeates the elderly population, affecting over 10% of adults older than 65^1^. The National Institute on Aging and Alzheimer’s Association now classifies AD as a continuum of biomarker and neuroimaging levels under a biological construct^2^, indicating that biology and cognitive decline are intertwined. Although many techniques are available to diagnose cognitive decline, undetected dementia remains at 55-68% globally^3^. Patients are often unaware of their cognitive decline^4^, limiting their ability to adequately address physical and mental limitations caused by dementia. Furthermore, 15-35% of patients older than 65 who are offered cognitive screening refuse to perform cognitive assessments, especially if they do not personally know anyone affected with AD^5,6^. Even after being referred by a community pharmacist to a physician for a follow-up cognitive study, almost 80% of pre-screened patients did not see a physician within 60 days, and over 40% of patients were unwilling to pay for additional cognitive screening^7^. Older adults often view cognitive assessments as embarrassing, invasive, and confusing^8,9^. Without a proper diagnosis, patients may postpone end-of-life planning until their memory further deteriorates, or they may be incapable of completing an advance directive (i.e., living will) if their memory has already deteriorated^10^. Therefore, it is imperative that cognitive function is accurately and affordably diagnosed early in disease progression in a manner that makes the patient feel comfortable.

The Clinical Dementia Rating (CDR)^11^ is a widely used cognitive diagnostic assessment^12^ that is comparable to the gold standard dementia diagnostic criteria^13^. The CDR examination requires 8-9 hours of training before it can be administered by a researcher or clinician (see https://knightadrc.wustl.edu/cdr/cdr.htm). The patient examination takes between 30-90 minutes^14^ and requires assessing differences in patient memory compared with the memory from a reliable informant (e.g., family member or caregiver)^15^. In contrast, a blood draw is less intrusive and could be completed in a matter of minutes by healthcare professionals without specific cognitive training. Therefore, a blood draw establishing a biological basis for cognitive decline would improve diagnostic accessibility and potentially increase patient response rate.

We used blood RNA levels in the Alzheimer’s Disease Neuroimaging Initiative (ADNI) dataset to predict patient CDR. We performed dimensionality reduction to avoid overfitting on the training dataset by using an analysis of variance (ANOVA) as an initial filter to extract the most significant microarray probes, similar to other machine learning analyses of microarray data^16^. We filtered 49,386 RNA probes based on their ANOVA p-values, and we used various p-value thresholds to assess predictive accuracy. Probes exceeding the p-value threshold were used as input features in a support vector machine to predict CDR scores for 741 participants in the ADNI dataset.

## Methods

Data used in the preparation of this article were obtained from the Alzheimer’s Disease Neuroimaging Initiative (ADNI) database (adni.loni.usc.edu). The ADNI was launched in 2003 as a public-private partnership, led by Principal Investigator Michael W. Weiner, MD. The primary goal of ADNI has been to test whether serial magnetic resonance imaging (MRI), positron emission tomography (PET), other biological markers, and clinical and neuropsychological assessment can be combined to measure the progression of mild cognitive impairment (MCI) and early Alzheimer’s disease (AD).

We used expression data from an Affymetrix HG U219 Array that cost less than $150 per array in 2015. ADNI preprocessed the raw expression values using the Robust Multi-chip Average (RMA) normalization method before mapping and annotating the probe sets to the hg19 human reference genome. The ADNI Genetics Core performed several other quality control measures on the dataset. Array plate randomization, gender and diagnosis balance, participant and probe quality control, and SNP-transcript cis-eQTL posterior probabilities were completed to ensure that analyses conducted on the dataset are not impacted by confounding factors. We also ensured that each individual had taken a CDR exam, which limited the available dataset to 49,386 probes across 741 participants whose cognitive abilities ranged from normal to severe dementia.

We labelled participants in one of three cognitive groups based on their most recent CDR score: cognitive normal (CDR=0), mild cognitive impairment (CDR=0.5), and probable AD (CDR≥1.0). We clustered CDR levels of 2.0 and 3.0 into the probable AD group to maintain predictive power because only 15 individuals had a CDR score of 2.0 and only one individual had a CDR score of 3.0. In total, 250 individuals were cognitive normal, 411 individuals had mild cognitive impairment, and 80 individuals had probable AD based on their CDR scores.

We conducted a one-way analysis of variance (ANOVA) on each of the 49,386 probes individually to test if expression levels significantly differ between the three groups. After a Bonferroni correction, our significance threshold was 1.012 × 10^−6^. We further assessed sex-specific biases in the three groups using the five X-inactive specific transcript (XIST) probes in the dataset, and we determined that no significant sex differences exist between the three cognitive groups (p-values= 0.0145, 0.017, 0.019, 0.041, and 0.068). We ordered all probes based on their ANOVA p-values from most significant to least significant and we identified significant probes. We then pruned our dataset based on the following alpha values: 1 × 10^−6^, 5 × 10^−6^, 1 × 10^−5^, 5 × 10^−5^, 1 × 10^−4^, 5 × 10^−4^, 0.001, 0.005, 0.01, 0.05, 0.1, 0.5, 1.0. These cutoff criteria were used to assess the predictive potential of combining RNA probes as input in a machine learning model. We used the Waikato Environment for Knowledge Analysis (Weka)^17^ implementation of sequential minimal optimization, which is a fast heuristic of a support vector machine. Support vector machines are non-probabilistic binary linear classifiers that separate training data so that the separator creates the largest gap possible between different groups. We scaled our input features in two ways: 1. Normalizing input features by rescaling the data between 0 and 1, and 2. Standardizing input features based on standard deviations from the sample mean. All other hyperparameters were left at their default settings in Weka. We performed 10-fold cross validation for each alpha value and scaled input values, repeating each analysis 10 times by randomizing our data for each replicate to minimize potential training split biases. We then assessed the predictive accuracy of each partition. Figure 1 depicts the process used to analyze the data.

**Figure 1:**
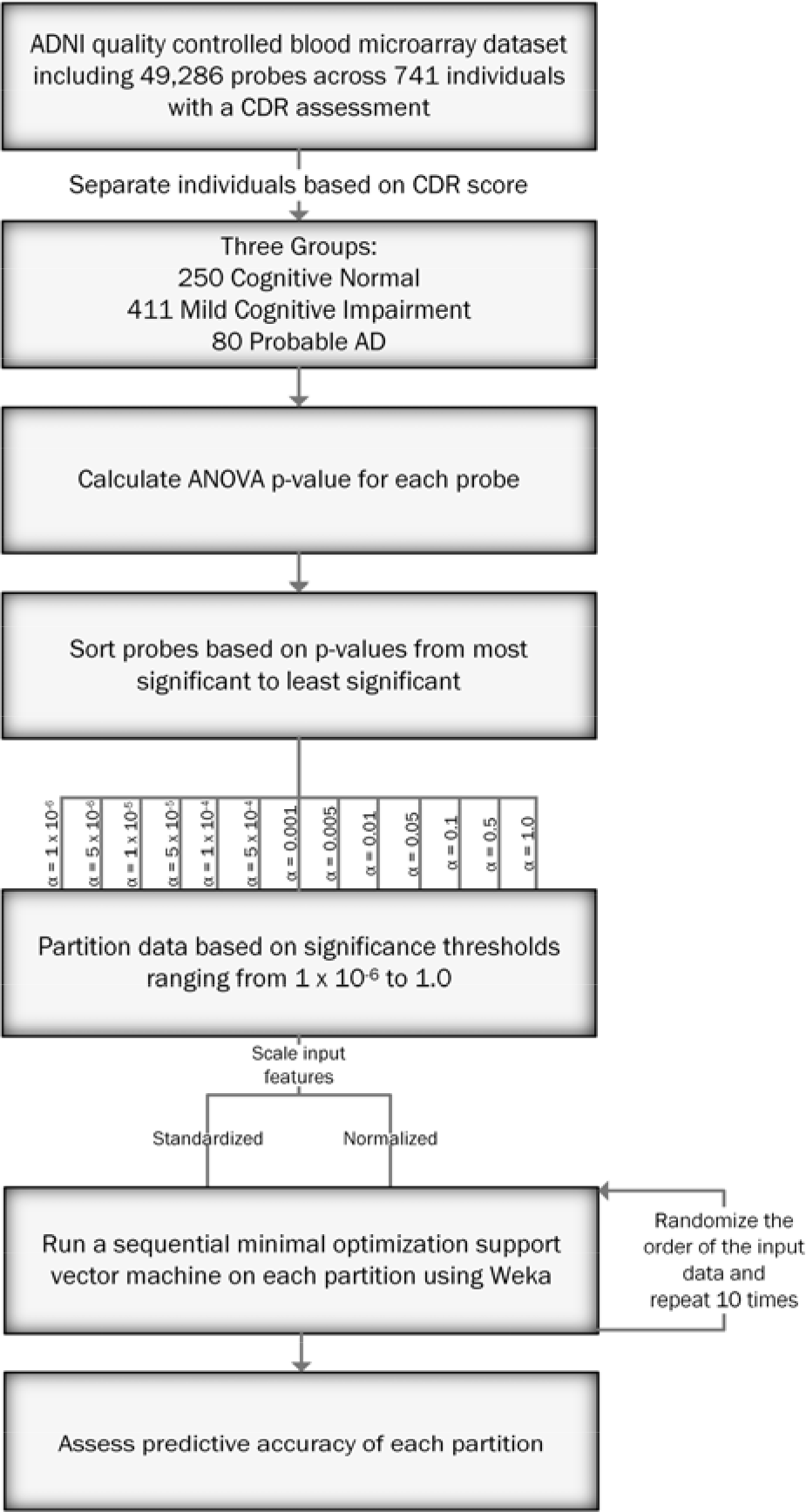
Flowchart depicting the analysis process to predict cognitive status using the ADNI blood microarray dataset.

## Results

We first analyzed each probe individually to determine if significant differences in RNA levels exist between the three cognitive groups. All three Chloride Intracellular Channel 1 (CLIC1) probes were in the top five most significant probes for the dataset, with one probe exceeding the Bonferroni threshold for significance. Table 1 shows the mean expression levels for the significant CLIC1 probe (11757474_x_at). The mean levels for the cognitive normal and mild cognitive impairment groups do −7 not significantly differ (p-value=0.30). However, the probable AD group has a significantly higher mean expression than the other two groups (p-value= 5.2781×10^−7^).

**Table 1:**
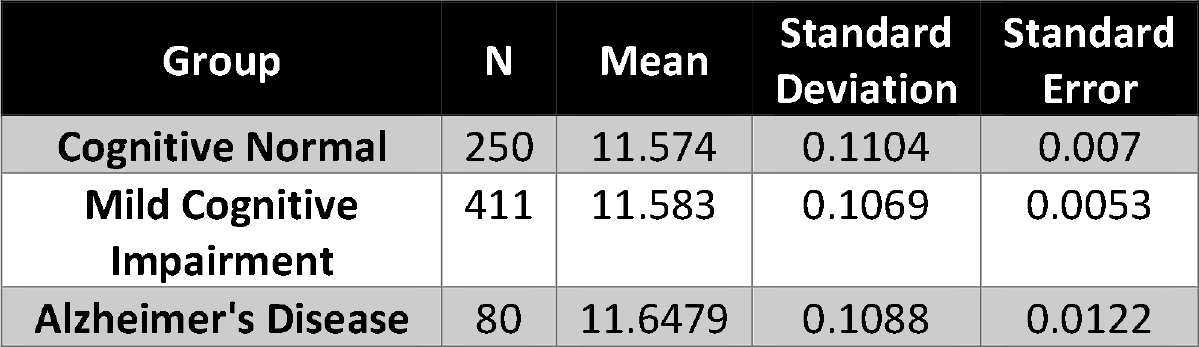
Mean RNA expression levels for the 11757474_x_at probe. The three groups are based on Clinical Dementia Rating (CDR): cognitive normal (CDR=0), mild cognitive impairment (CDR=0.5), and Alzheimer’s disease (CDR≥1.0). The mean RNA levels for the Alzheimer’s disease group significantly differs from the other two groups.

| Group | N | Mean | Standard Deviation | Standard Error |
| --- | --- | --- | --- | --- |
| Cognitive Normal | 250 | 11.574 | 0.1104 | 0.007 |
| Mild Cognitive Impairment | 411 | 11.583 | 0.1069 | 0.0053 |
| Alzheimer's Disease | 80 | 11.6479 | 0.1088 | 0.0122 |

Although mean RNA levels for CLIC1 probe 11757474_x_at statistically differ from mean RNA levels in the other two cognitive groups, that probe alone is insufficient to determine cognitive status. Therefore, we examined combinations of individually non-significant probes to predict CDR levels. Collectively, the probes roughly followed expected significance values, with about 5% of the probes having p-values less than or equal to 0.05. Figure 2 shows the ANOVA p-values for each probe and the Bonferroni corrected alpha value for the dataset.

**Figure 2:**
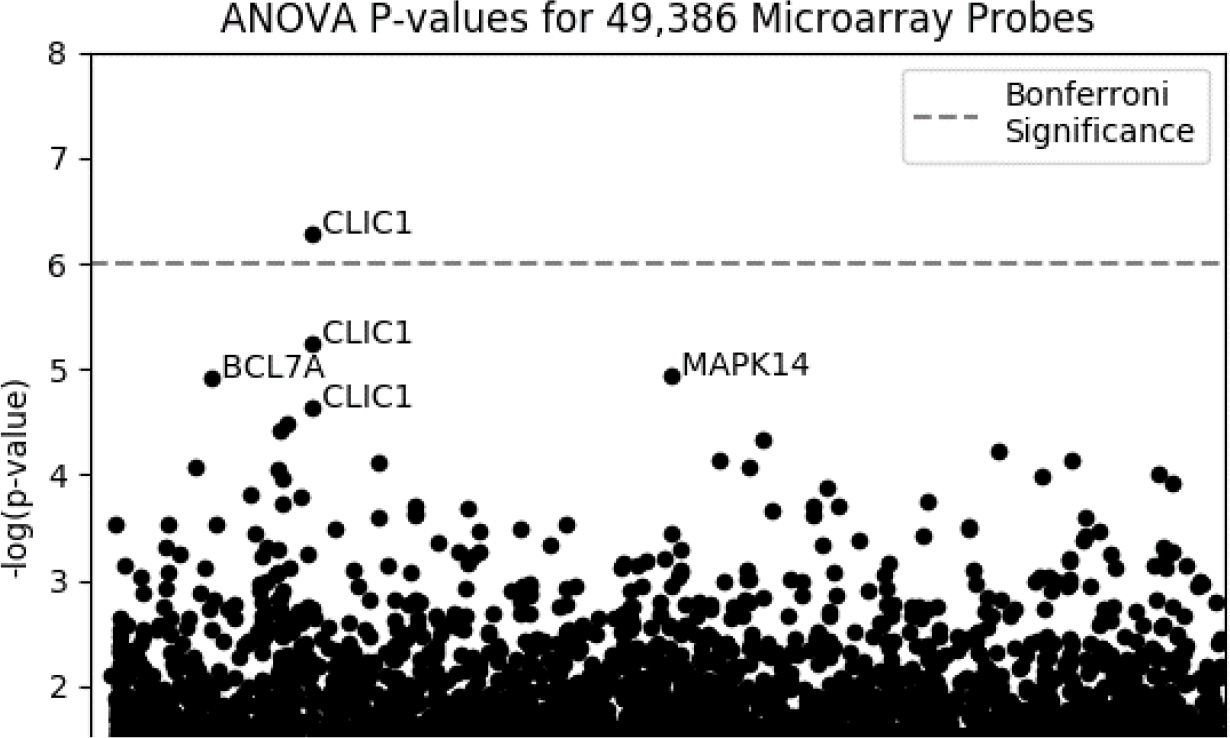
Negative log-transformed p-values for each probe, sorted by the gene name. The dashed line shows the threshold for significance after correcting for multiple tests. Only one probe for the Chloride Intracellular Channel 1 (CLIC1) exceeded the significance threshold.

We used 13 alpha values to assess the combined predictive potential of individually non-significant probes. Probes with p-values less than or equal to the selected alpha value were either standardized or normalized for two separate analyses. We created 10 permutated input files for the probes to calculate a standard deviation for the support vector machine on each alpha value. Figure 3 shows the percent accuracies and standard deviations in predicting cognitive status for each alpha value using 10-fold cross validation. The highest predictive power occurred when using an alpha of 0.1. The standardized permutations had a mean accuracy of 87.87 (s = 1.02)%, while the normalized permutations had a mean accuracy of 87.25 (s = 0.77)%. A t-test showed a significant difference between the maximum percent accuracies between the normalized and standardized datasets (p-value= 5.039 × 10^−38^), although the difference in the mean predictive accuracy was minimal (0.52%). All ten permutations in both datasets had a 0% false positive rate for AD. Incorrect predictions for AD patients were 2.91 to 3.25 times more likely to be classified as mild cognitive impairment than as cognitive normal in the normalized and standardized datasets, respectfully. Similarly, incorrect predictions for cognitive normal individuals were 4.51 to 4.81 times more likely to be classified as mild cognitive impairment than as AD in the normalized and standardized datasets, respectfully. Using additional probes with higher p-values in the model significantly decreased the predictive accuracy. Including all probes in the support vector machine lowered the predictive accuracy to the baseline, 55.46%.

**Figure 3:**
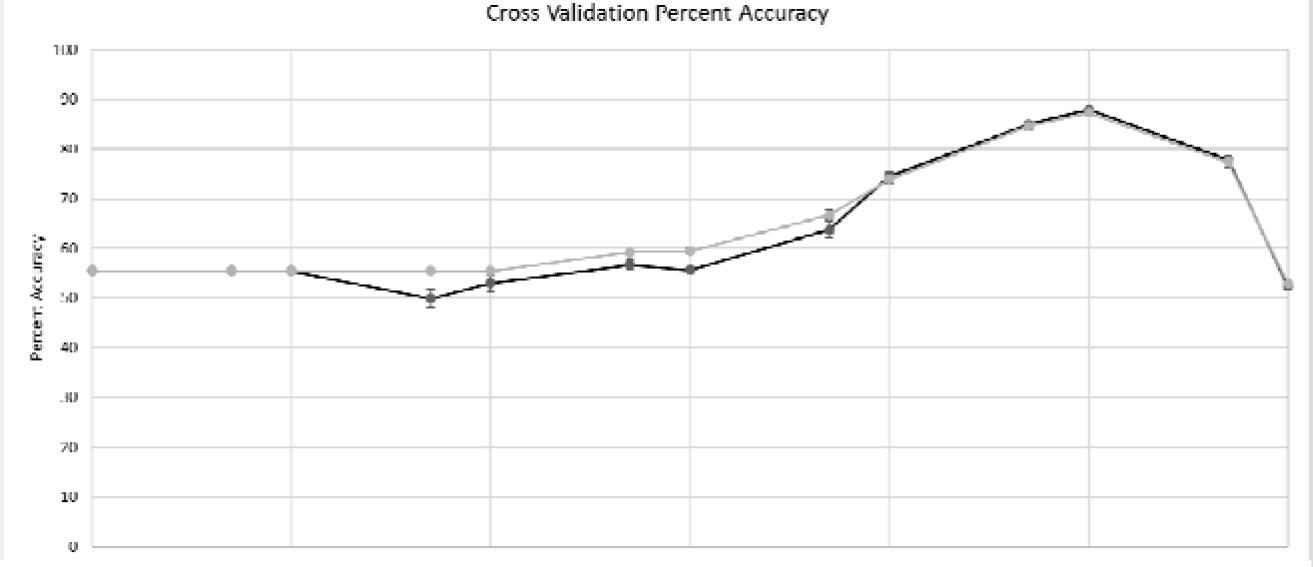
Percent Accuracy for the Support Vector Machines. Accuracies for predictions using both the standardized and normalized datasets are plotted.

## Discussion

We identified one significant probe in the chloride intracellular channel 1 (CLIC1). CLIC1 has previously been linked to AD and induces neurotoxin production in the presence of β-amyloid (Aβ) protein^18^. A direct link between CLIC1 expression and Aβ-induced microglial activation has also been established^19^. Our analyses show that significantly higher levels of CLIC1 exist in AD patients compared with cognitive normal and mild cognitive impairment groups. These results support previous indications that CLICL1 levels increase in AD patients. They also show that these differences are detectable in peripheral blood. However, CLIC1 expression alone is insufficient to accurately diagnose cognitive status in an individual.

Our analyses indicate that RNA levels from a blood microarray can accurately indicate cognitive decline in an individual. Although most RNA levels do not significantly differ between cognitive groups, combining the small differences in RNA levels in individually nonsignificant probes significantly increases the predictive accuracy of the model. We show that we can accurately predict CDR using a support vector machine on blood RNA levels. The support vector machine was able to increase the predictive accuracy from a 55% baseline to almost 90%. There was also a clear directionality in the predictions, with incorrect predictions for cognitive normal and AD patients more likely to be one cognitive group away instead of two cognitive groups away from the diagnosis. This directionality in predictions indicates that blood RNA levels gradually change as a patient progresses from a cognitive normal state to AD.

Our analyses also suggest that combining nonsignificant traits that suggest an association (e.g., p-value less than 0.1) may increase the accuracy of disease assessments. At the population level, low body mass index^20^, vital exhaustion^21^, changes in retinal microvasculature^22^, and many other risk factors each indicate early signs of Alzheimer’s disease. However, the natural variance within the population prevents each of these methods from being used as a diagnostic technique. Similarly, individual RNA probes within the ADNI dataset have levels that significantly overlap between cognitive groups and cannot be used in isolation to diagnose a patient. However, predictions became highly accurate when considering thousands of minor differences in RNA levels after eliminating excess noise in the dataset. Therefore, using a similar technique to combine other input features (e.g., body mass index, psychological health, retinal vessel density, etc.) in machine learning models may further increase the accuracy of early AD diagnoses.

## Data Availability

Data used in this study are publicly available through the Alzheimer's Disease Neuroimaging Initiative.

## Acknowledgements

We appreciate the contributions of Brigham Young University and the Office of Research Computing at Brigham Young University for supporting our research and our facilities.

Data collection and sharing for this project was funded by the Alzheimer’s Disease Neuroimaging Initiative (ADNI) (National Institutes of Health Grant U01 AG024904) and DOD ADNI (Department of Defense award number W81XWH-12-2-0012). ADNI is funded by the National Institute on Aging, the National Institute of Biomedical Imaging and Bioengineering, and through generous contributions from the following: AbbVie, Alzheimer’s Association; Alzheimer’s Drug Discovery Foundation; Araclon Biotech; BioClinica, Inc.; Biogen; Bristol-Myers Squibb Company; CereSpir, Inc.; Cogstate; Eisai Inc.; Elan Pharmaceuticals, Inc.; Eli Lilly and Company; EuroImmun; F. Hoffmann-La Roche Ltd and its affiliated company Genentech, Inc.; Fujirebio; GE Healthcare; IXICO Ltd.; Janssen Alzheimer Immunotherapy Research & Development, LLC.; Johnson & Johnson Pharmaceutical Research & Development LLC.; Lumosity; Lundbeck; Merck & Co., Inc.; Meso Scale Diagnostics, LLC.; NeuroRx Research; Neurotrack Technologies; Novartis Pharmaceuticals Corporation; Pfizer Inc.; Piramal Imaging; Servier; Takeda Pharmaceutical Company; and Transition Therapeutics. The Canadian Institutes of Health Research is providing funds to support ADNI clinical sites in Canada. Private sector contributions are facilitated by the Foundation for the National Institutes of Health (www.fnih.org). The grantee organization is the Northern California Institute for Research and Education, and the study is coordinated by the Alzheimer’s Therapeutic Research Institute at the University of Southern California. ADNI data are disseminated by the Laboratory for Neuro Imaging at the University of Southern California.

